# Development and evaluation of rapid data-enabled access to routine clinical information to enhance early recruitment to the national clinical platform trial of COVID-19 community treatments

**DOI:** 10.1101/2021.01.15.21249724

**Authors:** Caroline Cake, Emma Ogburn, Heather Pinches, Garry Coleman, David Seymour, Fran Woodard, Sinduja Manohar, Marjia Monsur, Martin Landray, Gaynor Dalton, Andrew D Morris, Patrick F. Chinnery, UK COVID-19 National Core Studies Consortium, FD Richard Hobbs, Christopher Butler

**Affiliations:** Health Data Research UK, Gibbs Building, 215 Euston Road, London, NW1 2BE; NHS Digital, Skipton House, 80 London Rd, Elephant and Castle, London SE1 6LH; NHS DigiTrials, Skipton House, 80 London Rd, Elephant and Castle, London; DHSC, Department for Health and Social Care, 39 Victoria St, Westminster, London SW1H 0EU; Nuffield Department of Primary Care Health Sciences, University of Oxford, Gibson Building 1st Floor, Radcliffe Observatory Quarter, Woodstock Road, Oxford, OX2 6GG; HDR UK Oxford, Big Data Institute, Nuffield Department of Population Health, University of Oxford, Oxford OX3 7LF; Department of Clinical Neuroscience & Medical Research Council Mitochondrial Biology Unit, School of Clinical Medicine, University of Cambridge, Cambridge CB2 0QQ, UK

**Keywords:** COVID-19, data, healthcare, clinical trials, public and patient involvement and engagement, recruitment, primary care

## Abstract

**Background:** The COVID-19 pandemic has presented unique challenges for rapidly designing, initiating, and delivering therapeutic clinical trials. PRINCIPLE is the UK national platform investigating repurposed therapies for COVID-19 treatment of older people in the community at high risk of complications. Standard methods of patient recruitment were failing to meet the required pace and scale of enrolment.

This paper describes the development and appraisal of a near real-time, data-driven, ethical approach for enhancing recruitment in community care by contacting people with a recent COVID-19 positive test result from the central NHS Test and Trace service within 24-48 hours of their test result.

**Methods:** A multi-disciplinary team was formed to solve the technical, ethical, public perception, logistical and information governance issues required to provide a near-real time (within 24-48 hours of receiving a positive test) feed of potential trial participants from test result data to the research team. PRINCIPLE was also given unique access to the Summary Care Record (SCR) to ensure safe prescribing, and to enable the trial team to quickly and safely bring consented patients into the trial. A survey of the public was used to understand public perceptions of the use of test data for this proposed methodology.

**Results:** Prior to establishing the data service, PRINCIPLE registered on average 87 participants per week. This increased by up to 87 additional people registered per week from the test data, contributing to an increase from 1,013 recruits to PRINCIPLE at the start of October 2020 to 2,802 recruits by 20^th^ December 2020.

While procedural caveats were identified by the public consultation, out of 2,639 people contacted by PRINCIPLE following a positive test result, no one raised a concern about being approached.

**Conclusions:** This paper describes a novel approach to using near-real time NHS operational data to recruit community-based patients within a few days of presentation with acute illness.

This approach increased recruitment, and reduced time between positive test and randomisation, allowing more rapid evaluation of treatments and increased safety for participants. End-to-end public and patient involvement in the design of the approach provided evidence to inform information governance decisions.

**Trial registration:** PRINCIPLE is funded by UK Research and Innovation and the Department of Health and Social Care through the National Institute for Health Research.

EudraCT number: 2020-001209-22

ISRCTN registry: ISRCTN86534580

REC number: 20/SC/058

IRAS number: 281958

## BACKGROUND

By the 26^th^ November 2020, over 1.6 million people in the UK had tested positive for COVID-19, and 57,896 people had died within 28 days of a positive COVID test result^1^. There were 62 million cases worldwide and 1.45 million deaths^2^. There are currently no specific treatments with proven effectiveness suitable for use in the community against COVID-19.

The Platform Randomised trial of INterventions against COVID-19 In older people (PRINCIPLE)^3^ was established in March 2020 as an Urgent Public Health, UK-wide National Priority Platform that aims to evaluate the effects of repurposed drugs for people in the community with SARS-CoV-2 infection who are at high risk of complications. The study currently aims to evaluate whether repurposed medicines speed recovery and reduce the need for hospitalisation and reduce deaths in people with suspected or proven COVID-19. The trial design allows trial arms to be stopped for proven effectiveness, futility or safety reasons, and for arms to be replaced or added. At inception, PRINCIPLE used traditional methods of primary care recruitment through general practices that were opened as study sites, with primary care clinicians checking eligibility and dispensing or prescribing study medications themselves.

Between 2^nd^ April 2020 and 4^th^ October 2020, PRINCIPLE registered 1,630 people who were identified as eligible at screening. Of these, 1,013 patients were randomised (or ‘recruited’) over those first 6 months. It was important to develop new approaches that would accelerate recruitment and produce results that would inform care more rapidly.

Although the total number of people hospitalised with COVID has been small relative to the number who have remained in community settings (circa 200,000 hospitalised, more than 1.3 million with a positive test result have remained in community settings in the UK), prior to this project initiating, PRINCIPLE had recruited far fewer compared with the number recruited to RECOVERY (the national clinical trial to identify treatments that may be beneficial for people hospitalised with suspected or confirmed COVID-19).

Recruitment into community based COVID-19 Randomised Control Trials (RCTs) has been limited by: initially having to set up several hundreds of general medical practices as sites (each with a small patient pool); challenges in dispensing and getting study medications to participants; efficiently confirming eligibility and safe prescribing in the trial; and, discussing possible trial participation with potentially eligible people in a timely way. These steps added time delays to the process that resulted in many potential participants feeling either recovered or being admitted to hospital by the time eligibility was confirmed.

Pre-COVID-19, routine health care data have been used very effectively to identify and recruit eligible participants with common, non-communicable diseases to clinical trials^4^ and observational cohort studies^5^.

With major limitations on access to general practices due to the pandemic, PRINCIPLE recruitment was expanded in July 2020 to include a centralised approach option, where participants could join the trial online or with telephone support, with online consent and central eligibly checks based on data received from participants primary care clinicians. Furthermore, an option was added to enable study medications and materials to be couriered to participants homes by the research team rather than relying on primary care clinicians.

The aim of this project was to develop an effective and trusted approach to rapidly increase the number of recruits (from a total of 1,013 recruits to 3,000 recruits) from the community in the UK during the autumn/winter 2020/21 by using near-real time testing data and for the clinical study team to access NHS Summary Care Records to support safe prescribing.

This paper describes an approach for identifying potentially eligible participants for RCTs, within 24 hours of receiving a positive SARS-COV-19 test result, that is achieving important numbers of additional recruits to PRINCIPLE. This approach overcomes some of the challenges of clinical trial recruitment in distributed settings.

The approach also adds to the collective evidence of patient experience of recruitment using routine healthcare data and also outlines a model for the future delivery of clinical trials - an approach being developed by the NHS DigiTrials health data research hub.

## METHODS

A multi-disciplinary team was convened by the UK National Core Studies Programmes (Therapeutics; Data and Connectivity) to solve the technical, ethical, public perception, logistical and information governance questions required to use near-real time data to recruit clinical trial participants.

The initiative was a partnership between the PRINCIPLE investigators and coordinating centre at the University of Oxford, the UK National Core Studies Programmes (Therapeutics; Data and Connectivity), the NHS Digital Information Governance Team, the Information Commissioner’s Office, NHS Test and Trace, the NHS DigiTrials Health Data Research Hub, the Department of Health and Social Care (DHSC), and the HDR UK Public & Patient Involvement and Engagement Team. An overview of the key programmes and organisations involved is provided in Figure 1.

**Figure 1:**
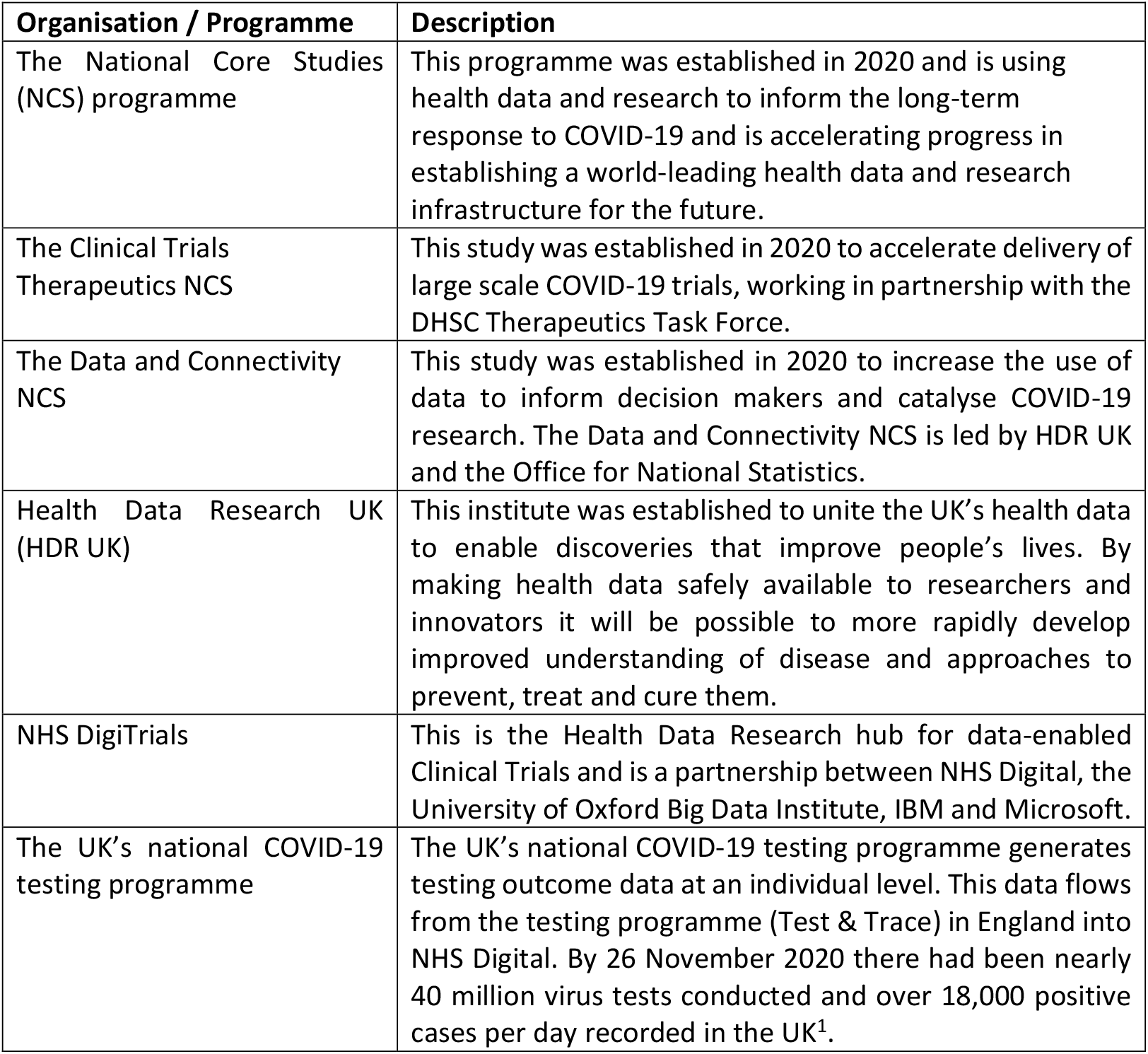
Overview of Programmes and Organisations involved.

The team followed a rapid, agile project approach to rapidly define requirements, design the technical approach, identify rate limiting steps and problem solve.

The first challenge was to determine whether it was feasible for positive test results to be received by the trial team within 24-48 hours of the recipient receiving the result. Any longer would reduce the likelihood of the participant being suitable for the trial (as they may have deteriorated or be approaching recovery by then). The NHS DigiTrials team mapped the data flow and timing from the point of test receipt to the time the data was received within NHS Digital. An overview process map is shown in Figure 2.

**Figure 2:**
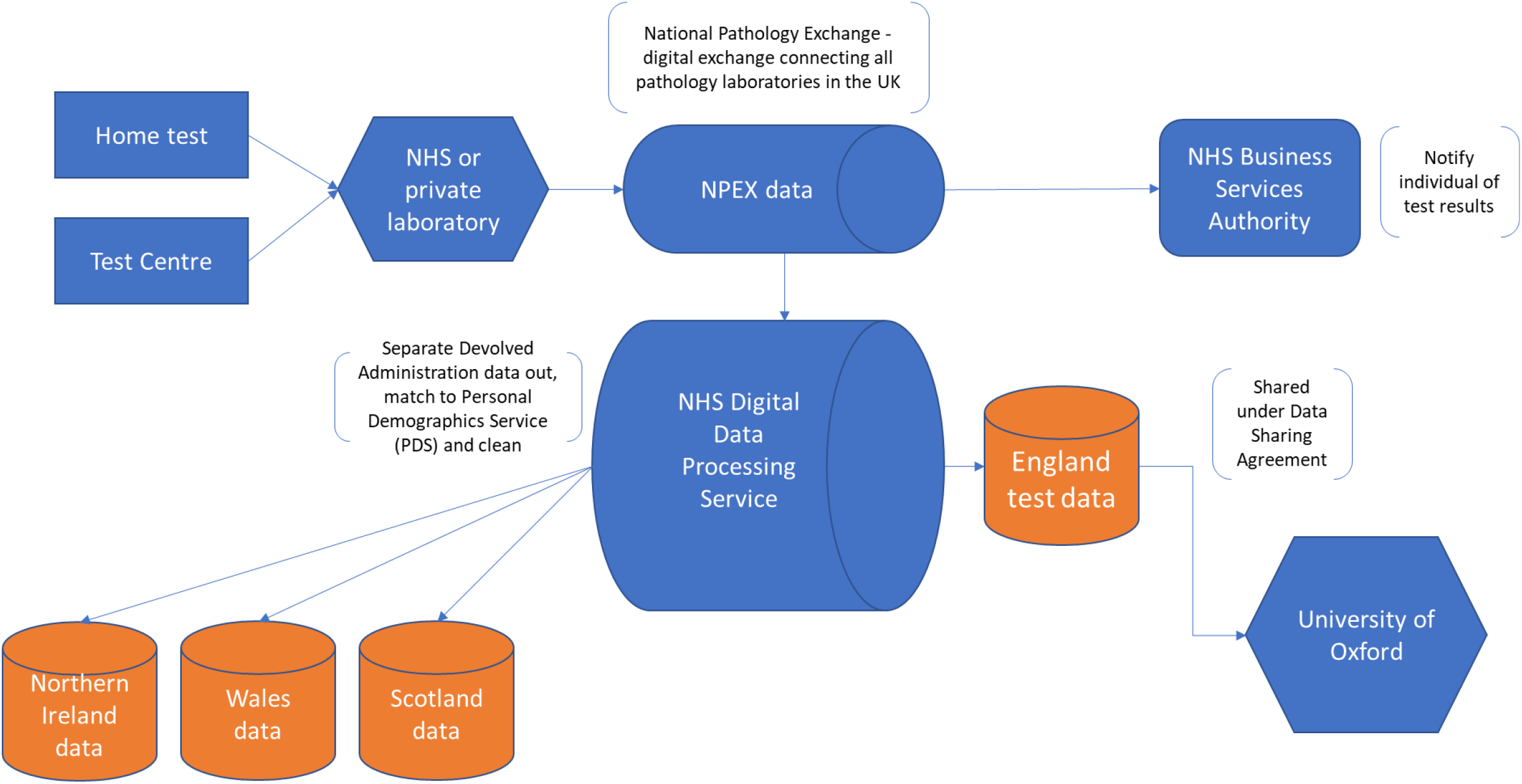
Overview Process Diagram for test data route to the clinical trial.

During the COVID-19 pandemic, the Secretary of State for Health and Social Care issued NHS Digital with a Notice under Regulation 3(4) of the National Health Service (Control of Patient Information Regulations) 2002 (COPI) to require NHS Digital to share confidential patient information with organisations entitled to process this under COPI for COVID-19 purposes.

The test result data needed to be sent to PRINCIPLE as identifiable data so that they could verify eligibility and contact eligible candidates. The eligibility criteria for the Trial are described in Figure 3. As identifiable data was involved, this required a review of the data sharing and data protection requirements. The Information Commissioner’s Office (ICO) team and NHS Digital Information Governance teams were engaged to review whether the actions constituted direct marketing, whether Privacy and Electronic Communications Regulations (PECR) applied, and clarifications around privacy notices, data sharing and data controllership.

**Figure 3:** PRINCIPLE Trial eligibility criteria. *Anyone experiencing COVID-19 symptoms, which started within the last 14 days may be eligible to join the PRINCIPLE trial if they are aged over 65; or over 50 with an underlying health condition*. From https://www.principletrial.org/ accessed on 29 December 2020

The NHS Digital Information Governance reviews concluded that in the context of a global pandemic an updated privacy notice on the DHSC and NHS Digital websites was the minimum requirement to enable re-contact of people with positive test results. Furthermore, it was important to ensure that the clinical trials team contacted potential candidates after the individual had received the positive test result.

The team also applied for access to the Summary Care Record (SCR), to ensure safe prescribing to participants who presented to the trial team. This enabled the team to effectively and efficiently verify the information provided by the individual and to ensure safe prescribing of trial medication.

Prior to the work described in this paper, NHS Digital had not yet shared Test result data with any clinical trials organisation for recruitment purposes. Use of the SCR had been limited to direct care purposes and research and clinical trials were specifically excluded. Therefore, the information governance implications and the public and patient perceptions of doing this were unclear and untested. The SCR agreed access approach and requirements are set out in Figure 4.

**Figure 4:** Summary Care Record Access. SCR is a live database and is accessed via Role-Based Access Controls (RBAC) through smartcards. For a patient who has provided consent to join the trial and agreed for their SCR to be accessed, the data will be accessed by a clinician to support safe prescribing decisions immediately prior to prescribing medications. A Summary Care Record will only ever be accessed on one occasion for each individual participant. The recommendation from the SCR team is to create a new role under (RTH ODS code) Oxford University Hospitals specifically for the trial to access SCR. This allows access to SCR via SCRa/Spine Portal and does not include emergency access. RBAC rule allow for SCR access to be time limited for the duration of the trial. The application to access SCR to support safe prescribing in the context of this clinical trial is considered to be a secondary use. For this reason SCR access was added to the existing data sharing agreement with the trial which provides a wrapper of contractual controls. It is recognised that this arrangement goes against the existing agreed policy for SCR which has been limited to direct care purposes only. However, this exception was agreed recognising the high priority of the trial, the compelling reasons put forward by the trial and the very specific set of circumstances of the Covid-19 pandemic. This included the fact that the trial is using commonly used drugs where allergies and interactions known and common (i.e. not a novel therapeutic), the need for timely recruitment within a specific clinical pathway and the data contained in the SCR is suitable to meet this purpose.

In parallel, a survey with members of the public was developed to understand public perceptions of the proposed approach. Survey respondents were provided the following scenario for context only: *A UK Wide National Priority Clinical Trial, endorsed by all UK Chief Medical Officers, being run by the University of Oxford, is exploring a possible treatments for COVID-19 in the community, and is looking for participants to join within 24-48 hours of receiving a positive COVID-19 test. In particular, they are looking for people 50 years and over to join this clinical trial*.

Each of the issues raised through the governance and the patient and public survey was reviewed and addressed in the design of the data release and recruitment approach.

The PRINCIPLE team contacted eligible candidates by phone and signposted them to the trial. The approach was tested through batches of data for people aged 50 – 75 years from NHS Digital to the PRINCIPLE Trial team. Once the data flows were tested, the PRINCIPLE Trial received contact data on 200 people per day aged 50 years plus with positive test results from 13th November 2020.

The majority of the 50 years plus age range didn’t meet the trial criteria as they didn’t have the required co-morbidities so the data export age range was changed to 65-75 years on 24^th^ November.

Initially, people were contacted by phone to ask whether they would be interested in participating in the trial. From the 29^th^ November, the method was changed slightly to also offer support to the participant through the recruitment process during the same call, resulting in enrolment into the trial.

From 26^th^ November PRINCIPLE began to receive 300 contacts a day aged 65-75 years from NHS Digital and from 4^th^ December PRINCIPLE received 400 contacts a day.

The tested approach was rolled out with tracked measures of efficiency, effectiveness and patients were asked to provide comments on their experience of the approach.

## RESULTS

Mapping the positive test result data flow from Test and Trace to NHS Digital showed that data could be made available to the Trial team within 24-48 hours of an individual receiving a positive test result.

Prior to establishing this new data service, the PRINCIPLE trial was registering on average 87 people per week. The number of people registered over the period from 16^th^ November to 20^th^ December increased from 3,080 to 4,092. The registration rate increased from up to 208 people per week to 325 people per week. Over this 5-week period, up to 87 additional people per week were registered from NHSD, accounting for up to 31% of the weekly registrations. The cumulative registrations are shown in Figure 5.

**Figure 5:**
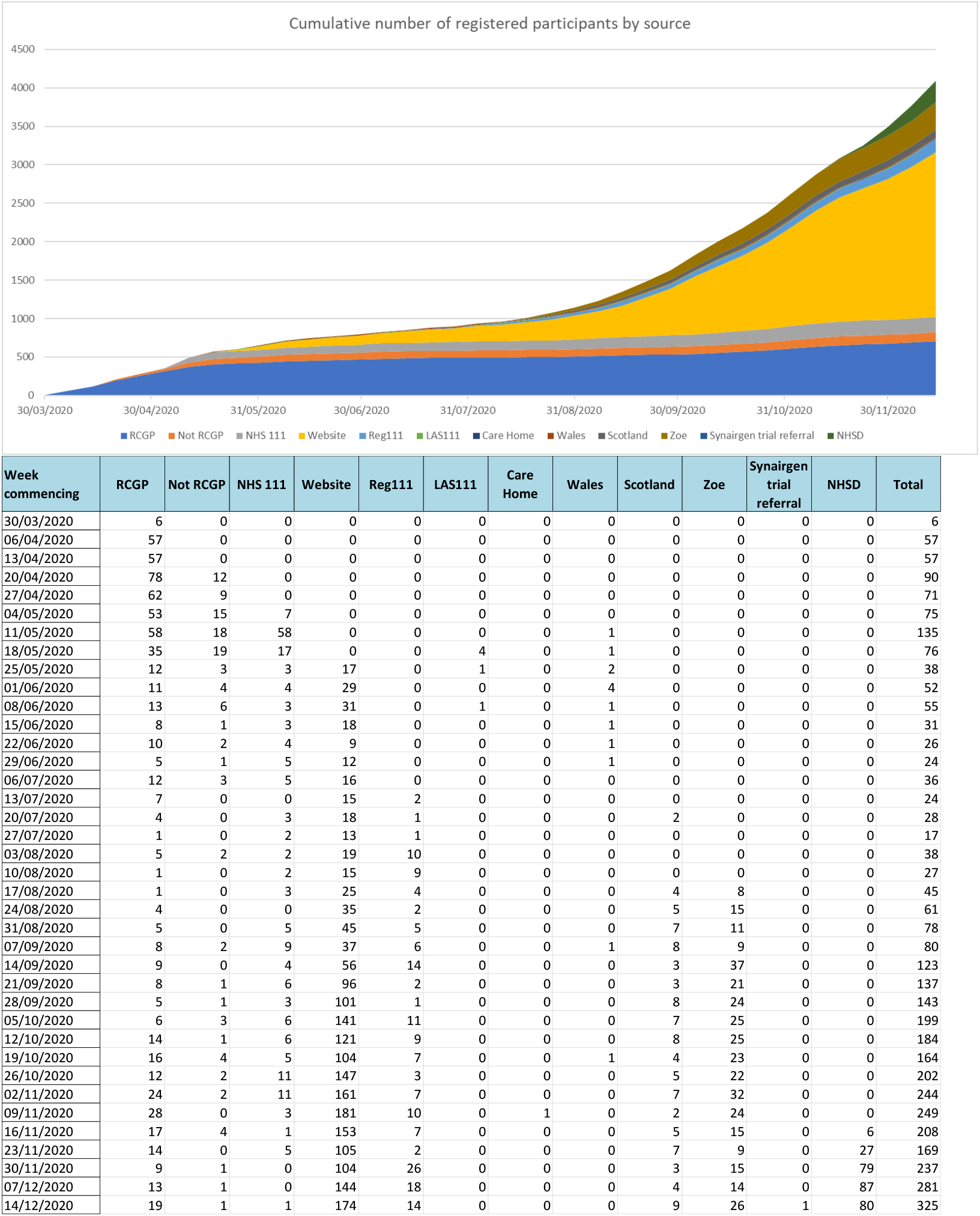
Number of people registered into PRINCIPLE.

Some registered patients are subsequently identified as not eligible and are not randomised for recruitment. The number of people recruited to PRINCIPLE increased from 1,521 at the start of November to 2,802 by 20^th^ December. The recruitment numbers are shown in Figure 6.

**Figure 6:**
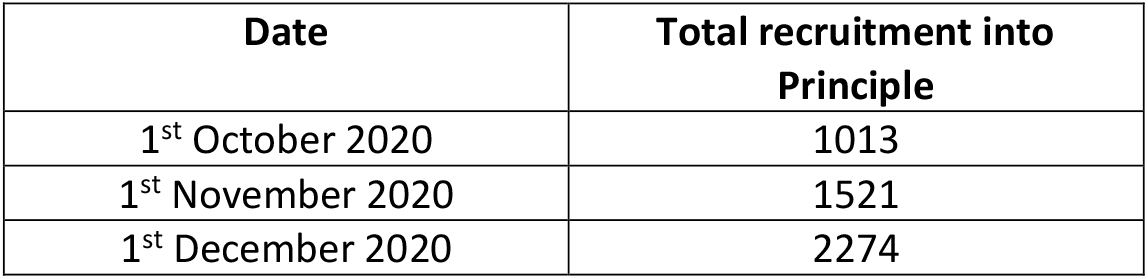
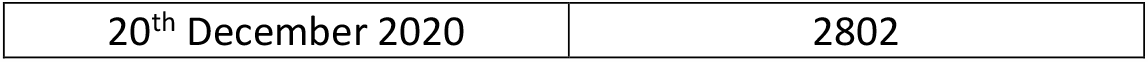
Recruitment into PRINCIPLE, number of people.

Figure 7 shows the recruitment call outcomes for people contacted using the test data. 731 people were contacted between 13th November and 24th November 2020, prior to starting the enrolment support. 4 people were recruited (0.5%). The majority did not answer their phone (297) and 123 people requested for information to be emailed to them. 1,908 people were contacted between 25th November and 4^th^ December 2020 and received the enrolment support. 87 people were recruited (4.6%). The total number of calls made between the 13^th^ November - 4^th^ December is 2,639 which has yielded 91 participants recruited over the phone and a further 326 requesting to receive trial information via email. GPs were then contacted for each individual deciding to participate to verify whether they would be suitable.

**Figure 7:**
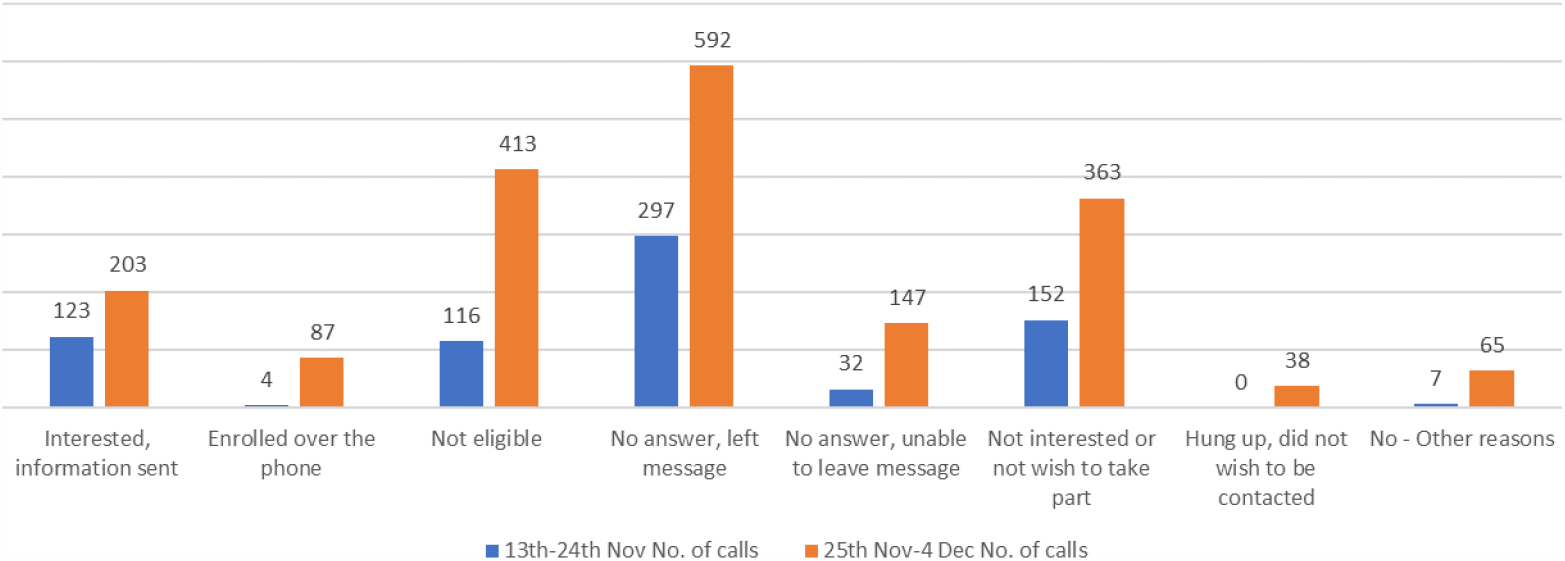
Call outcomes (for people identified via the NHSD test data) *After 24*^*th*^ *November, age range of contacts changed from 50+ to 65-75 years*

Access to the SCR was received on 3^rd^ December 2020 and became fully operational on the 8^th^ of December. Prior to using the SCR, it was taking the trial team from 1 to 10 days to confirm participant eligibility by securing the necessary participant data from the participant’s GP. This time was predominantly associated with efforts to contact busy GP surgeries.

Use of the SCR has now enabled the process of confirming participant eligibility by the trial team to be more efficient – requiring less input from participant GPs, reducing the number of study recruiters required in the trial team, hence making it a more efficient process. SCR access has led to safe prescribing checks reducing from 1-10 days per participant down to just 30 minutes per participant. This reduction in time from screen to confirming eligibility is critical given the urgency of recruiting people soon after diagnosis.

Through established patient and public panels and networks at HDR UK 92 people provided responses to the patient and public survey within 5 days. This was conducted in advance of establishing the service to understand what 92 people think they and others would feel about this approach.

97% of respondents believed this would be an ethical use of data and assuming all concerns were addressed, respondents were comfortable with receiving the message from the Oxford University team running the clinical trial (46%) or NHS (51%).

As Figure 8 shows, whilst 29% were very uncomfortable, 68% of people would be comfortable or very comfortable with a test result and email/text/phone contact data being passed on to researchers so that they could be invited to consider participating in a national priority UK clinical trial of treatments for COVID-19.

**Figure 8:**
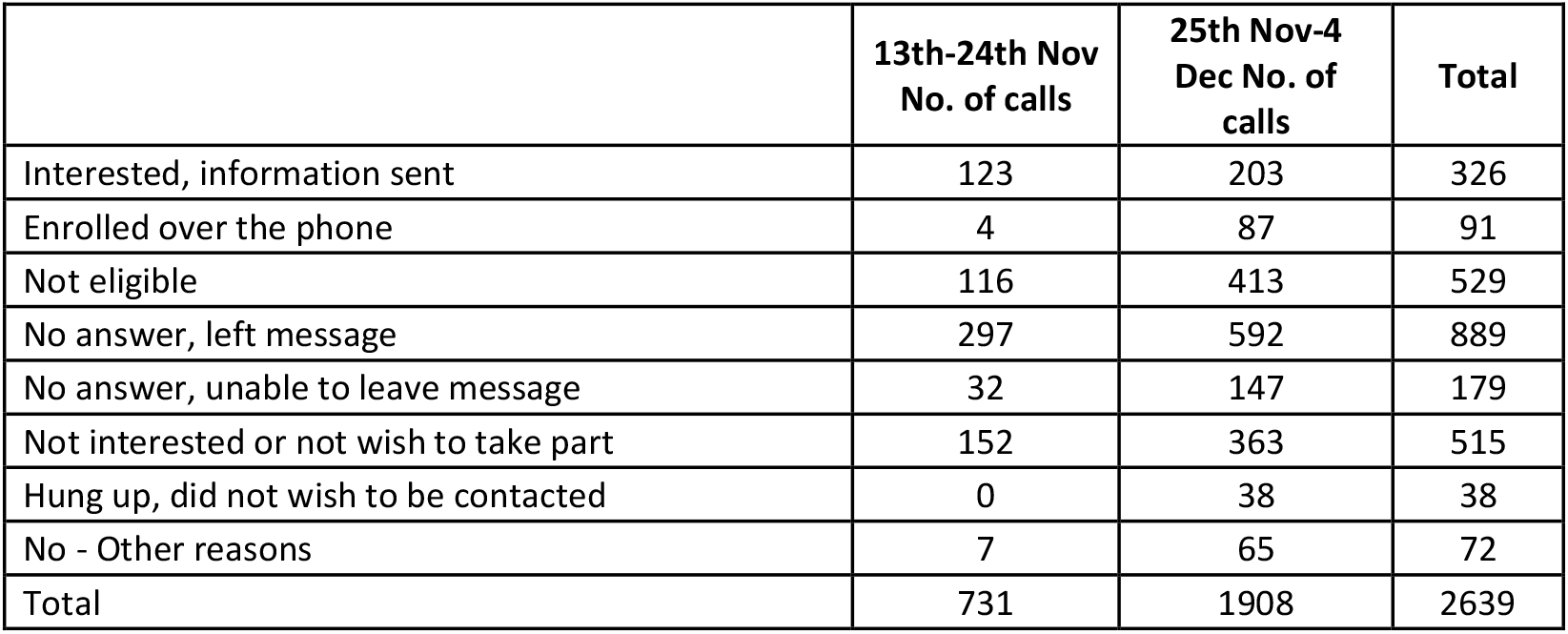
Patient and public survey responses to the question: If you had tested positive for COVID-19, and you were aged 50 and over, how comfortable would you feel?

Only 45% would be comfortable or very comfortable with receiving an automatic invite by email or text within 24 hours of test result inviting them to consider joining a clinical trial of possible treatments for COVID-19 illness, whereas 54% were uncomfortable or very uncomfortable.

Figure 9 shows that they would be most comfortable with receiving the invitation by email (81% were comfortable) and less comfortable with phone (23% were comfortable) or text (46% were comfortable). In particular, they would be least comfortable with receiving the invitation via the NHS Test and Trace app^6^ (4%). The sequence of actions was important to responders, as they felt that the individual needed to receive their test result, and appropriate information, guidance and support before being invited to participate in a clinical trial.

**Figure 9:**
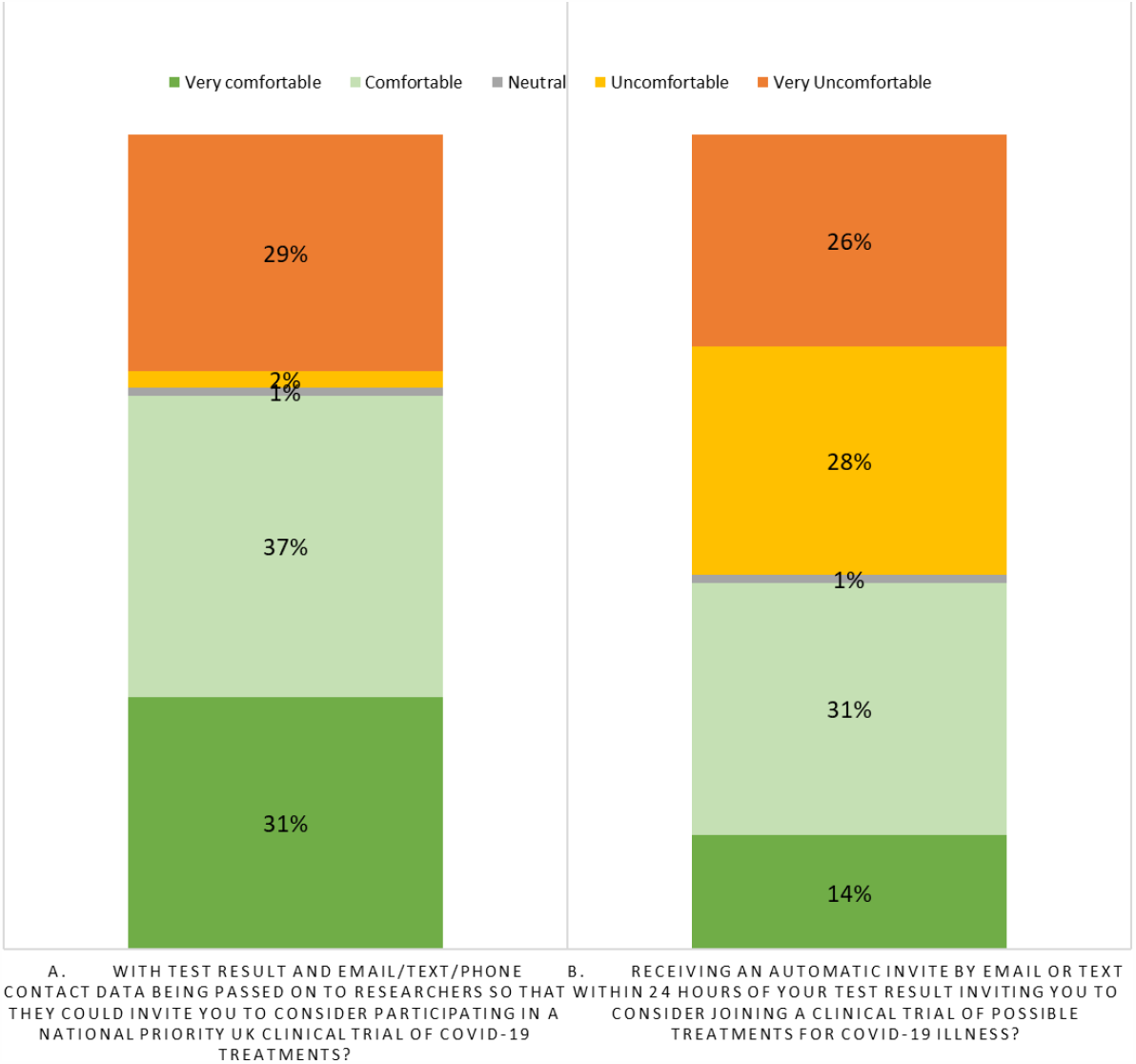
Patient and public survey responses to the question: Given the speed at which this invite needs to be sent (i.e., soon after a COVID-19 positive test result), would you feel comfortable with receiving an invite?

The full set of survey responses, including free text comments, is provided in Figure 10. As well as providing insights on public perceptions, the patient and public survey also demonstrated an efficient process to rapidly inform the team’s understanding and test an approach with members of the public through the HDR UK Patient and Public Networks.

**Figure 10:**
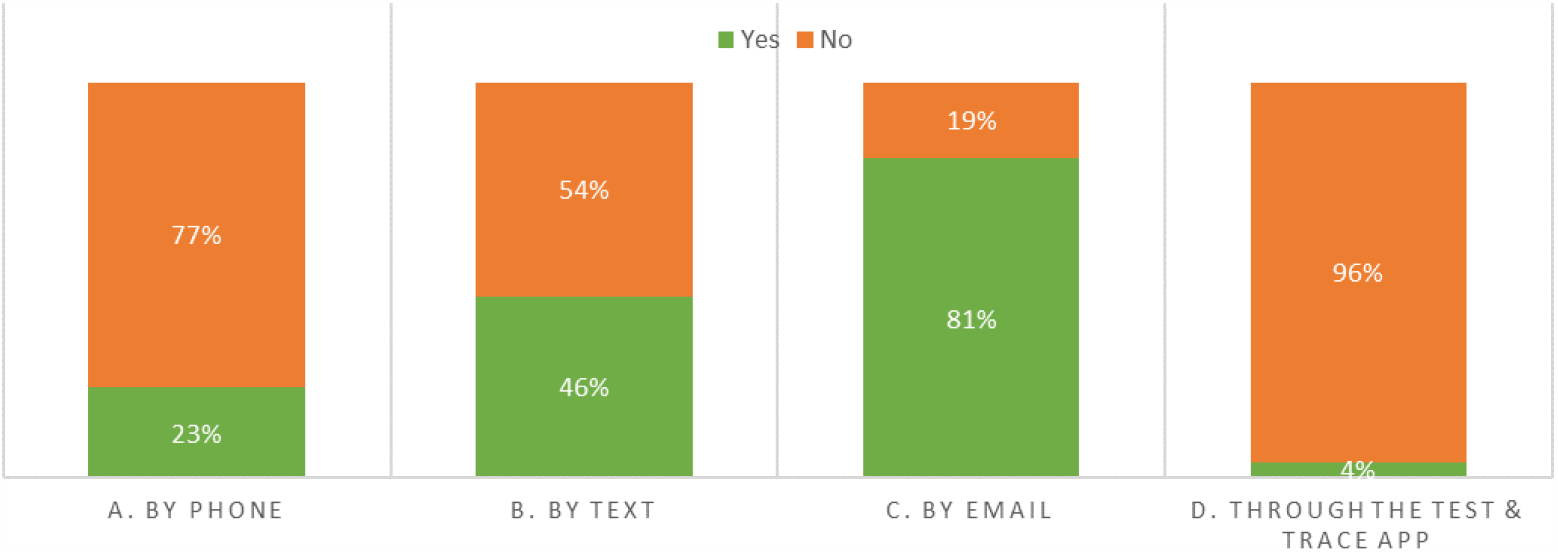
Patient and Public Perception Survey Responses. **•92 responses from Patient / Public Voices** **•Survey respondents were given the following scenario and were told this scenario is to provide context only** *A UK Wide National Priority Clinical Trial, endorsed by all UK Chief Medical Officers, being run by the University of Oxford, is exploring a possible treatments for COVID-19 in the community, and is looking for participants to join within 24-48 hours of receiving a positive COVID-19 test. In particular, they are looking for people 50 years and over to join this clinical trial*.

The Trial team decided to use phone calls to contact the participants for many reasons including experience in other primary care trials where texts/emails has yielded fewer recruits than phone calls or face to face trial discussion; and sending texts or emails to potential participants removes the real-time engagement between study team member and the participant. They took into account the concerns raised through the patient and public survey about telephone contact and crafted a script to specifically address each of the issues.

Patient experience was recorded for all people contacted and this showed that 38 people out of the 2639 (1.4%) hung up whilst the trial was being explained (Figure 7). Furthermore, 0 people raised concerns about being approached following a positive test result.

## DISCUSSION

Recruiting to the national priority community COVID trial using the additional mechanisms of study team access to test data and Summary Care Record data was associated with increased recruitment of people with positive COVID-19 test results in the community setting. The registration rate increased from up to 208 people per week to 325 people per week, with up to 87 people per week from the NHSD test result route, accounting for up to 31% of registrations per week. Furthermore, although the patient and public survey, which was conducted prior to establishing the service, indicated that 31% of people would be uncomfortable or very uncomfortable about results being passed onto researchers for this purpose, there were no concerns raised by any of the 2639 people contacted during the actual service.

The recruitment of people in secondary care settings can be organised by the hospitals enabling recruitment to secondary care clinical trials to be significantly higher for secondary care participants compared with primary care participants, despite there being many more people with COVID-19 in primary rather than secondary settings (circa 200,000 hospitalised with COVID-19, whereas more than 1.3 million have remained in community settings in the UK). As a comparison on 28^th^ November 2020 RECOVERY had recruited 19,000 participants, approximately 10% of all patients hospitalised with COVID-19, whereas PRINCIPLE had recruited 2,224 participants, approximately 0.15% of all patients diagnosed in the community (however, not all patients with a positive test in the community is at risk of getting complications or within the PRINCIPLE criteria age range).

The more rapid identification of people eligible for the PRINCIPLE trial means that more people are being given the opportunity to contribute to trials of potential treatments that may reduce deterioration and the need for secondary care treatment. Increased recruitment will also allow more interventions to be evaluated in the trial, with shorter time to results.

Trial clinical team access to the SCR enhances efficiency and safe prescribing. However, the SCR service, prior to this trial, had been limited to direct care only with research purposes specifically excluded. The Trial needed to demonstrate a clear and compelling use case for accessing this data in the current pandemic which was considered on its merits including input from an independent panel of experts. Access was allowed under the pandemic COPI powers as secondary use. The SCR webpages were updated to ensure transparency in relation to the access granted.

Further work is required to evaluate which aspects can be replicated for recruitment into clinical trials for other primary care treatments. This work was conducted with English test and with SCR data, further work is also required to extend this approach across all four UK nations.

The integration of the patient and public survey in the design of the data-enabled recruitment workflow, and the measurement of patient experience during the roll-out of the approach, provided important information to team members designing the information governance process and the sequencing of contacting people. This approach demonstrates the effectiveness and efficiency of patient and public involvement and engagement through organised groups such as the HDR UK Patient and Public Network and can be built on and developed further to better inform future clinical trials.

## CONCLUSION

We proposed and then demonstrated a new way of enhancing early recruitment to a clinical trial in a community setting, via access to SARS-COV-2 test result data and the use of SCR data to ensure efficient and safe prescribing. Potential treatments could therefore be initiated earlier in suitable patients who consented, also shortening the time to results. Although effective COVID vaccines were licensed from December 2020, it is still anticipated that it will be several months before these are fully rolled out, and longer for many parts of the world. Furthermore, despite highly effective vaccines, there will be many people who continue to require effective treatments for COVID-19. Therefore, the faster effective treatments can be found, affected people may be able to recover quickly and be hospitalised less often. This novel and efficient approach to an acute condition trial may be an important new methodology for acute incident studies in primary care, which are traditionally more difficult to recruit to than trials in long term conditions.

## Data Availability

All data generated or analysed during this study are included in this published article

## LIST OF ABBREVIATIONS

COPI: Control of Patient Information Regulations
COVID-19: 2019 novel coronavirus
DHSC: Department for Health and Social Care
GP: General Practitioner
HDR UK: Health Data Research UK
ICO: Information Commissioner’s Office
LAS: London Ambulance Service
NCS: National Core Study
NHS: National Health Service
NHS: 111 NHS urgent medical help service
NHSD: NHS Digital
PECR: Privacy and Electronic Communications Regulations
PRINCIPLE: Platform Randomised Control Trial of Interventions against COVID-19 in older people
RCGP: Royal College of General Practitioners
RCT: Randomised Control Trial
RECOVERY: Randomised Evaluation of COVID-19 Therapy
SCR: Summary Care Record

## DECLARATIONS

### Ethics approval and consent to participate

IRAS PROJECT ID 281958, REC Reference 20/SC/0158

### Consent for publication

Not applicable

### Availability of data and materials

All data generated or analysed during this study are included in this published article

## Authors’ contributions

SM analysed and interpreted the public survey data. EO analysed and interpreted the recruitment data. CC formed and coordinated the multi-disciplinary study team and was a major contributor in writing the manuscript. All authors read and approved the final manuscript.

## Acknowledgements

The authors gratefully acknowledge the contributions of all individuals who provided their input into this work, including to all 1030 GP practices who are supporting the PRINCIPLE trial and the 92 volunteers who provided their views and insights in the patient and public survey.

## FIGURES, TABLES AND ADDITIONAL FILES

Survey responses

**Question 1:** If you had tested positive for COVID-19, and you were aged 50 and over, how comfortable would you feel:

|  | Very comfortable | Comfortable | Neutral | Uncomfortable | Very Uncomfortable |
| --- | --- | --- | --- | --- | --- |
| a. With test result and email/text/phone contact data being passed on to researchers so that they could invite you to consider participating in a national priority UK clinical trial of COVID-19 treatments? | 31% | 37% | 1% | 2% | 29% |
| b. Receiving an automatic invite by email or text within 24 hours of your test result inviting you to consider joining a clinical trial of possible treatments for COVID-19 illness? | 14% | 31% | 1% | 28% | 26% |

**Question 2:** Do you believe this would be an ethical use of data?

☒ Yes – 97%
☒ No – 3%

**Question 3:** Does this access and use of data cause any concerns or lead to any questions *(open text)*?

- I think it is the acquisition of your results in the first place that is more problematic than their eventual use, as you can consent to being involved in a trial but <u>your results are passed on with no consent</u>.
- I think it breaches GDPR unless overridden by government emergency legislation
- How will you ensure you won’t be contacting patients who have opted out of taking part in research? Particularly if those people have changed their minds?
- Depending on severity of infection, would not want to have to deal with this as there would be other priorities
- What about patients in care and residential homes who don’t have the capacity to consent to being contacted?
- Worries about personally targeted with restrictive interventions.
- Need to be careful as you may actually contact people who pass away from COVID or an unrelated illness. If this became public knowledge it could discourage people from attending a COVID test.
- There needs to be consideration that these patients and their families maybe stressed and upset.
- People do not like their details being passed onto third parties without their permission.
- It could backfire and discourage people from using the NHS app.
- An automated email can be explained easier and means that you are hopefully not passing patient information on.
- Who exactly has access to this data and how is it stored? Who exactly is this data being shared with? How long is this data being stored for? On which data base is this information being stored on? Will there be a named contact to adjust and amend personal data?
- How data is stored? What happens to it long term?
- For use I would want to know the pedigree of research.
- In terms of access It would be better as part of the test procedure if you consented to your results being passed into a repository where only ‘approved’ researchers could get it.
- The principle of using the data in this way is certainly acceptable I think, especially considering the scale of the pandemic and the urgency of handling data in this scenario. That would not excuse misuse of data (such as passing it on for commercial use)

**Question 4:** If all concerns were addressed, would you be comfortable with receiving the message from (*please tick all that apply*):

|  |  |
| --- | --- |
| The Oxford University team running the clinical trial | 46% |
| NHS | 51% |
| Another organisation -please state the type of organisation:<br>- NIHR<br>- GP rather than just NHS | 3% |

Additional comments:

- Would prefer one point of contact
- Would accept an email from the NHS as I then wouldn’t feel that my details have been passed onto another party. You need to think about patient confidentiality.
- As contact information would be given to NHS it would be reasonable for them to contact as they have already had to with the results of the COVID test. Also one’s GP = part of the NHS – has this contact information. However, if it was another team contacting me (such as the trial team) I would be very concerned
- The various controversies swirling round this appalling government’s handling of the pandemic, notably the allocation of contracts to non-NHS bodies, does not inspire confidence. The NHS label just about still inspires confidence
- I would feel less happy about third party involvement
- As long as it was flagged that the data was going to be shared with the people getting in touch with you. I would be angry if I got a text from another organisation and hadn’t been told they would be contacting me/seeing/accessing my data
- What credentials would make me trust it’s the Oxford University team?
- All formats for contact are acceptable but, if it were from a non-NHS source, I would want to see sort of official statement from the NHS saying they endorsed it.
- Verification as to who it was from with contact details.
- Whoever makes the contact need some additional assurance about my data being used in the limited ways needed for the specific research, and keep clear lines of communication in the event that I wanted to follow up on anything about the research and use of my data.

**Question 5:** Given the speed at which this invite needs to be sent (i.e. soon after a COVID-19 positive test result), would you feel comfortable receiving an invite:

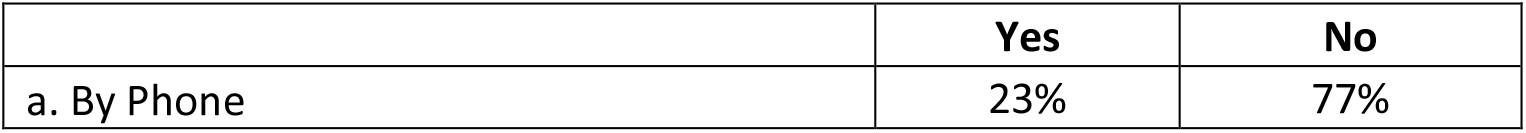
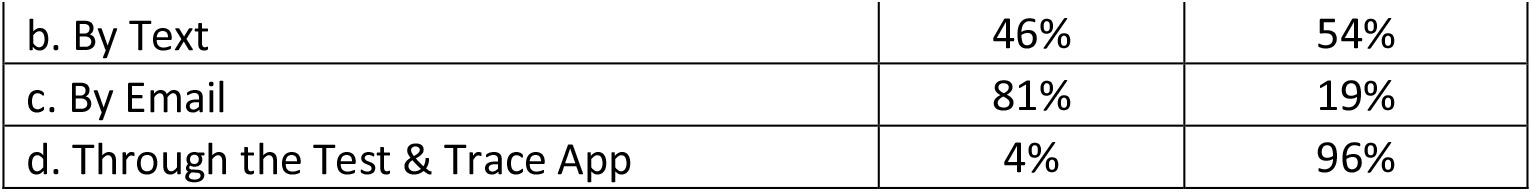

**Additional comments for**

**By phone**

- Provided it was backed up by an email with background info and someone I could ask questions of after reading the info.
- I would certainly be very cautious of a telephone call that did not show the caller number or did not have a regular number. They would have to give me some unique information about myself that would only be known to a genuine caller, but this would probably not be possible for GDPR reasons.
- There are too many phone scams going on at the moment so either the phone would not be answered as it is from an unknown number or there would be a lot of trust issues
- Just to add that we get so many problem phone, email and SMS, despite various filters, that I am immediately suspicious of any I don’t recognise or am not expecting. Most are easily recognisable for what they are, but a few are very plausible hence I am extremely cautious with them
- I am happy with text and email because I should be more able to tell if it is genuine but on the phone - could be from anyone.
- I would need some form of re-assurance that the approach was genuine as I am always suspicious of unsolicited approaches.
- Phone calls can be intrusive and I tend not to answer calls from unknown callers
- I do not have a “smart” phone and hearing loss makes use of landline difficult

**By Text:**

- I would only be happy for a text to prompt me to go to another source for detailed info. I would not say ‘yes’ to a text.
- If you had to incorporate all this info to someone it would be a long text. Perhaps in a text just put the necessary info and ask them to confirm agreement to a follow up phone call where the individual had to contact a known NHS number that they could google to verify.
- Just because a patient has a mobile you can’t assume that the person with COVID has the capacity to consent to take part in a research study. It could be an older person with a mobile used only for contacting family in an emergency.

**By email:**

- Email would be best as it’s the easiest way to keep a hard copy of the information.
- Text messages could be used to alert to an email invitation if needed for speed

**Through the test and trace app:**

- Given the poor uptake of the test and trace app I would suggest not
- Test & Trace and “the app” have been contaminated by Govt’s blind faith in private sector and antipathy to public service delivery of public health. This is exacerbated by the leadership of Test & Trace
- Test and trace is failing to get up to speed. Set a time limit after which, unfortunately, my participation will no longer be useful.
- A lot of issues with the app are coming up in the various PPI groups and networks I have but also stories in the media. The public don’t trust the app so could then not trust you if that’s how you communicate
- Do not trust this app (esp. after data being shared with police) and do not believe it works
- Lots of people are either not downloading it or have downloaded and then deleted it for various reasons so not reliable
- I wouldn’t choose the app because I assume it wouldn’t give me the direct link to the researchers (I’m assuming here that the others could) and I’m not sure I’d trust it in terms of keeping my data private.

**Any other format? If so, what format?**

- You will need to be careful about it looking legitimate – not a scam such as you might get in an email/text/phone. What about things like ‘my chart’ or platforms used by GP? I appreciate not everyone is on my chart, but you could do it via GP text service – we are all used to getting texts from GP

**Question 6:** What information would you want to see in this invite message?

- Clear identification of sender and reason as to why the invite was being sent
- Link to study format
- I would want details of the study and what I would be obliged to undergo
- Brief information about the study. Link or paper version for more information. Clear, easy to understand information about the trial so someone could say yes or no. I do think that a text/email should have enough info for the person to say yes or no but not too much so it’s overwhelming.
- There are a number of different trails around COVID and a great deal of confusion, I suspect the lay person does not realise how many. It would help if potential volunteers were given very specific details about the trial – i.e. is it to help develop a vaccine or a treatment regime to explain the differences from other trials.
- Contact details. Ethics. Safety. Why me? Patient friendly summary and information. Details of why I have been contacted.
- Links to pedigree of study. Details of what is involved. What are the potential risks? What are the potential benefits? A very clear consent statement. FAQs. How to opt out? How data is stored? What happens to it long term? I’d want clarity and assurances about how my data would be protected
- What information would you be able to give the patients about likely success and/or side effects? In participating, would you be ruling yourself out from the standard treatment methods if you get to that stage?
- Though I would be happy to be of use, I think it is important that potential participants should be aware that they are not obliged in any way to take part
- Clear comprehensive info about process and data security and the chance of being targeted by restrictive interventions.
- Most importantly, seeking my permission to participate and to use my own data, including a withdrawal option.
- Named contact for further information. Full details of the organisation sending it. The authority under which it has been sent, and the full rights the potential respondent has with regards to it.
- I’d like a named contact person in the research team, and a lay summary of what the research was about including its timelines and further intentions/scope for the researchers to be in contact with participants.
- The recipient may well be feeling ill or at least upset by having to isolate – this need this acknowledged first. The ethics of data collection will probably not be uppermost in their minds. To be too ‘objective’ can be off putting. Could learn something from the way blood donations work.
- Brief explanation, emphasis on speed, advice on how ill I am feeling and how this will affect my participation, what happens next, Contact details for further information, will they want participation from people I have been in contact with if they test positive too. Will home visits be involved, or travel?
- It would be good to get a yes/maybe and then a follow up call or email with all the info in so the person can be sure about what they are signing up for.
- Too much info in a text message/email is boring and people don’t read them/get put off by the length. Especially if there is technical language or jargon used which the NHS does a lot.
- Something like: now that you have tested positive you are being invited to join this trial: brief description of purpose of trial, what it involves for the individual, potential benefit to individual and others, acknowledgement of possible no/dysgenesis to individual. We understand that you will not be feeling well but the earlier you are enrolled into the trial the greater the value of your participation; discuss with next of kin and get back to us asap. Assurances about personal data security and the right to withdraw at any time. Contact details for more information or to answer any questions – needs to be contact details that come up on google to show it is genuine

**Question 7:** Would you want to know how your contact details have been sourced for this invite message e.g. *‘you have been contacted because you agreed for your data to be used for research purposes when you were tested for COVID-19’*?

☒ Yes – 92%
☒ No – 8%

**Question 8:** If ‘yes’, where you would expect to see this message?

- At the very start of the correspondence
- Up front in bold type in every communication - i.e. letter, survey, any report shared with participant.
- Should explain how they opted in, e.g. was it the Test and Trace app or when completing a form when doing the COVID-19 test
- If I’m not already aware of how my contact details have been sourced you should not be contacting me
- I would want to be reminded that I gave permission for this pretty early on
- There seems to be an important hiccough in the logical flow of communication from the research team here. At this stage (starting at Q 6), I would think I am still being contacted to give or confirm my permission for my data to be used, with an option to refuse. I don’t see where, at any prior point, I have expressly said ‘yes’ to this. So to receive a message, such as in the Q 7 example above, to say I’d already agreed, might upset me and lead me to withdraw. If I did that, I would also probably want a firm written assurance that my data had been fully removed from the trial. There needs to be an extra stage here, or before, where a recipient clearly gives consent (or not) to use their data, and based on the principles set out in Q 9 below.
- Don’t put it at the end or in the fine print, no one reads it and although you’re being ‘transparent’ it’s not the right way to do it because people don’t read it. It will be as though you found a loop hole instead.
- Sure, put it in the privacy notices but that’s not enough. It’s not transparent and feelings like you’re hiding it rather than being upfront when you get in touch with people.

Question 9: Would you need any reassurance that only non-commercial, publicly funded, ‘approved’ researchers are accessing the data, and sourced and used your data according to General Data Protection Regulations (GDPR)?

☒ Yes – 92%
☒ No – 8%

**Question 10:** If ‘yes’, where would you expect to see this reassurance and what would you want to know?

- This can go towards the end of the messages
- Link to confidentiality agreements
- On the first page of a summary or in the email. I don’t like this approach because patients could be opting in for their data to be used in multiple research studies in different countries. I’m currently opted out of research using data from my medical records due to a bad experience of a clinical trial so I’m probably biased by my concerns are genuine.
- I would expect a full list of all the approved researchers to be included in the email or letter.
- Back up info on web. How it meets all legal requirements, how data will be used, a reassurance that, if commercially successful, UK taxpayers, who have funded the initial NHS involvement, will share in any profits.
- Don’t really understand what it would mean but I assume it should be included
- Do not read those messages – only skim – but at least you’re covering yourself

**Question 11:** Any other comments? *(comments in italics came up <u>very</u> <u>frequently</u>)*

- *A positive test means the person will feel more like a patient and so probably wanting help/to help whereas general members of the public might be more concerned about being contacted in the first place in which case the data protection information might be of higher priority*
- *You need to go in this order*
  - *Test result*
  - *Confirm they have test result*
  - *Ensure they have information/guidance/support needed*
  - *Ask about participation in a clinical trial*
- *Understand speed is of the essence but you need to be a human first*
- *It seems to me that there is already a lot of resistance to requests for isolation when contact has been made with someone who has the virus, so contact and participation after contracting the virus might be equally as difficult. Making the process as personal and efficient as possible must be the best way to achieve a positive take up*.
- *Question around eligibility and what data you are sent to determine if I am eligible*
  - *Are you just getting my name, contact details and test result – if so, that’s fine*
  - *If you’re being sent my age, where I live, other medical conditions I have then I don’t trust it and you need to re-evaluate now*
- I would not want to make the decision at that time I suspect so either I think you should delay giving the invitation a few days or, if time is of the essence in deciding to participate, then I’m not sure I would participate
- If I was tired or feeling ill from COVID I would not be enthusiastic to be contacted by someone. Likewise, email gives a better degree of security and enables the individual to respond at a time when they feel able to do so.
- I am happy for commercial companies to be involved and, to a large extent, I think it would be necessary to bring any useful findings to bear on patient outcomes at scale and quickly.
- Whoever (inc NHS) is using the data I would expect to comply with all legal requirements inc GDPR
- There are a number of different trails around COVID and a great deal of confusion, I suspect the lay person does not realise how many. It would help if potential volunteers were given very specific details about the trial – i.e. is it to help develop a vaccine or a treatment regime to explain the differences from other trials.
- The desire for speed to introduce this must not overshadow the importance of protecting participant data and reassuring participants of the safety and integrity of the study.

